# Short-term Forecasting of Cumulative Confirmed Cases of Covid-19 Pandemic in Somalia

**DOI:** 10.1101/2020.06.18.20135053

**Authors:** Dahir Abdi Ali, Habshah Midi

## Abstract

Somalia has recorded the first confirmed Covid-19 case and first death case on March 16, and April 08, 2020, respectively. Since its arrival, it had infected 2,603 people and took the lives of 88 people while 577 patients were recovered as of 14 June, 2020. To fight this pandemic, the government requires to make the necessary plans accordingly. To plan effectively, the government needs to answer this question: what will be the effect of Covid-19 cases in the country? To answer this question accurately and objectively, forecasting the spread of confirmed Covid-19 cases will be vital. To this regard, this paper provides real times forecasts of Covid-19 cases employing Holt’s linear trend model without seasonality. Provided that the data employed is accurate and the past pattern of the disease will continue in the future, this model is powerful to produce real time forecasts in the future with some degree of uncertainty. With the help of these forecasts, the government can make evidence based decisions by utilizing the scarce resource available at its disposal.

## Introduction

Covid-19 pandemic is a communicable disease which resulted from the severe acute respiratory syndrome. Although it first started in December 2019 in Wuhan city, in China, it is currently world-wide epidemic as declared by World Health Organization ^1^. It currently infected more than 7.5 million people and killed more than 428,000 people globally ^2^ as of June 13,2020. Although the governments across the globe implemented some containment measures such as curfews, wearing masks, hand sanitization and so on to fight against this pandemic, the number of confirmed cases continued to show an increasing trend. While it imposed a serious threat to the public health sector, other sectors such as economies were also significantly affected ^3,4^.

Fever, shortness of breath as well as dry cough are the common symptoms of this Covid-19 pandemic^5^ while mild diarrhea, loss of smell, sore throat and muscle pain are among the uncommon signs of the novel coronavirus. The Covid-19 pandemic is transmitted through respiratory droplets which are generated when an individual sneezes or coughs. Also, a person can get infected by touching a contaminated surface or coming into a contact with an infected person ^6^. The maximum hours this virus can survive on the surface is up to 72 hours and the incubation period is on average five to six days. Nevertheless, it can reach up to 14 days.

Among the recommend measures that can be implemented to avert the spread of the Covid-19 infection are social distancing, hand sanitization, covering your nose and mouth when sneezing and coughing with a tissue or inner elbow and avoid touching your face with contaminated hands ^7^. In addition, health practitioners are strongly advised to wear face masks especially the surgical masks to protect themselves from contracting the virus. The gist of these recommendations are to keep the general public safe from Covid-19 pandemic.

Somalia has recorded the first confirmed Covid-19 case and first death case on March 16, and April 08, 2020, respectively. The government of Somalia has taken several measures to curb the spread of this virus and these include banning of local and international flights in and out of the country, closing all schools and learning institutions. In addition, the government imposed night curfew on April 13, 2020 to fight against this virus. In Somalia, Covid-19 had infected 2,603 people and took the lives of 88 people while 577 patients were recovered as of 14 June, 2020 ^8^.

Furthermore, the most hit-hard state is Benadir region, the capital city of Somalia, which recoded 1,300 total confirmed cases. To assist public health planning and effective policy making, forecasting Covid-19 cases became invaluable. To this end, this paper provides statistical forecasts for the cumulative confirmed cases of Covid-19 as well as the confirmed new cases using powerful time series model in the context of Somalia. These time series forecasts help us to better comprehend the current circumstances of Covid-19 cases and plan for the future.

## Data and methods

Data on Covid-19 confirmed cases from March 16, 2020 to June 13, 2020 were retrieved from Ministry of Health (MOH) [https://moh.nomadilab.org/]. The data set included the cumulative confirmed cases, new confirmed cases, total recoveries, total deaths and confirmed cases by gender and geographical location.

Simple time series forecasting techniques were employed to forecast the cumulative confirmed cases of Covid-19 pandemic. Models from exponential smoothing family were adopted to produce reliable forecasts ^9,10^. According to ^11,12^, these models provide forecasts with good precisions relative to its competitors especially when the sample size is small. These models have the capability of exposing different trend and seasonal patterns in time series forecasting. These patterns can either be additive or multiplicative or combinations of them. According to patterns seen in Fig1, we adapted Holt’s Linear trend model ^13^. Based on statistical model selection methods such as Akaike information criteria (AIC), we opted Holt’s Linear trend model with additive error and additive trend components.

## Results and Discussion

According to Fig1, the cumulative confirmed cases experienced an exponential pattern with increasing trend. The curve has not yet started to flatten or has not yet reached a plateau while a gradual surge was observed in number of recovered cases.

First strand of forecasts: From May 11 to May 20, 2020. We began our first set of forecasts at the end May 10, 2020 with thirty-three observations. The forecasted cumulative cases with 95% confidence intervals were produced at the end of May 10, 2020 are depicted in Table 1. The forecasted mean for cumulative confirmed cases 10-days-step was 1,521 with 95% prediction interval which ranged between 1,234 and 1,808 cases while the observed cumulative cases on May 20, 2020 was 1,573 cases. An absolute percent error of 3.3% was observed on May 20, 2020.

**Table 1.**
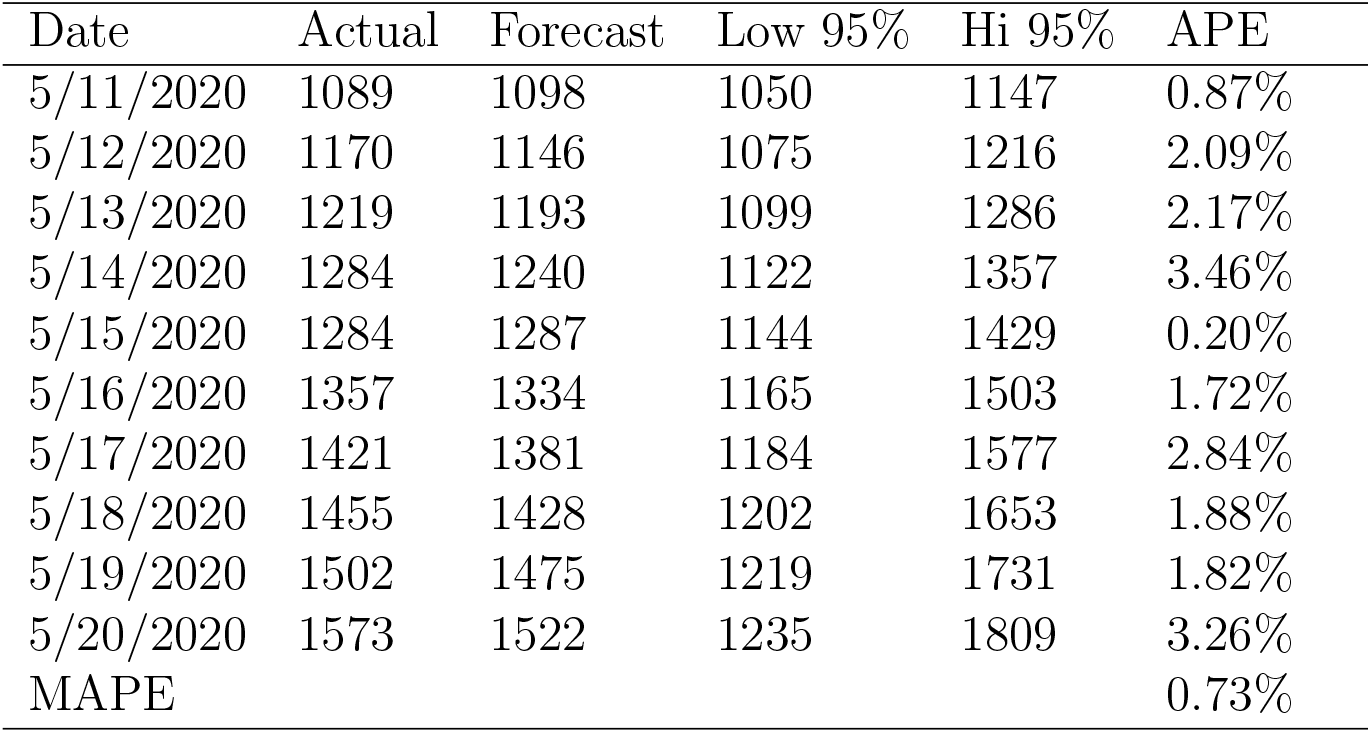
Forecasts, 95% prediction Interval and Absolute percent error.

| Date | Actual | Forecast | Low 95% | Hi 95% | APE |
| --- | --- | --- | --- | --- | --- |
| 5/11/2020 | 1089 | 1098 | 1050 | 1147 | 0.87% |
| 5/12/2020 | 1170 | 1146 | 1075 | 1216 | 2.09% |
| 5/13/2020 | 1219 | 1193 | 1099 | 1286 | 2.17% |
| 5/14/2020 | 1284 | 1240 | 1122 | 1357 | 3.46% |
| 5/15/2020 | 1284 | 1287 | 1144 | 1429 | 0.20% |
| 5/16/2020 | 1357 | 1334 | 1165 | 1503 | 1.72% |
| 5/17/2020 | 1421 | 1381 | 1184 | 1577 | 2.84% |
| 5/18/2020 | 1455 | 1428 | 1202 | 1653 | 1.88% |
| 5/19/2020 | 1502 | 1475 | 1219 | 1731 | 1.82% |
| 5/20/2020 | 1573 | 1522 | 1235 | 1809 | 3.26% |
| MAPE |  |  |  |  | 0.73% |

Nevertheless, the observed cumulative cases remained within the predication interval throughout this period. A mean absolute percent error of 0.73%, a measure of forecasting accuracy, was observed.

Second strand of forecasts: From May 21 to May 30, 2020. We updated our data set to include cumulative confirmed cases up to May 20, 2020 and again 10 days-step forecasts were generated. The mean estimates with 95% prediction intervals are shown in Table 2. The forecasted cumulative cases on May 30, 2020 was 2,090 cases with prediction interval of 1,836 and 2,345 cases while the realized cumulative confirmed cases stood at 1,909 cases. An absolute percent error of 9.5%(with the point forecast being slightly positively biased) was observed which is higher than the previous 10 days of forecasts. However, the forecasts still fall within the prediction interval.

**Table 2.** Forecasts, 95% prediction Interval and Absolute percent error.

| Date | Actual | Forecast | Low 95% | Hi 95% | APE |
| --- | --- | --- | --- | --- | --- |
| 5/21/2020 | 1594 | 1620 | 1572 | 1668 | 1.6% |
| 5/22/2020 | 1594 | 1672 | 1607 | 1738 | 4.9% |
| 5/23/2020 | 1594 | 1725 | 1640 | 1809 | 8.2% |
| 5/24/2020 | 1594 | 1777 | 1672 | 1882 | 11.5% |
| 5/25/2020 | 1689 | 1829 | 1702 | 1956 | 8.3% |
| 5/26/2020 | 1711 | 1882 | 1731 | 2032 | 10.0% |
| 5/27/2020 | 1724 | 1934 | 1759 | 2109 | 12.2% |
| 5/28/2020 | 1821 | 1986 | 1786 | 2186 | 9.1% |
| 5/29/2020 | 1821 | 2039 | 1812 | 2265 | 11.9% |
| 5/30/2020 | 1909 | 2091 | 1836 | 2345 | 9.5% |
| MAPE |  |  |  |  | 8.7% |

Third strand of forecasts: From May 31 to June 09, 2020. Again 10 days-step point forecasts and its 95 percent prediction intervals using data up until May 30,2020 are produced and presented in Table 3. The forecasted cumulative confirmed cases on June 09, 2020 were 2,331 cases while the actual confirmed cases were 2,409 cases. This time round, we recorded an absolute percent forecast error of 3.2 percent which was noticeably lower than previous forecasting error of 9.5 percent. This means that the forecasting uncertainty was higher in the preceding strand of forecasts.

**Table 3.**
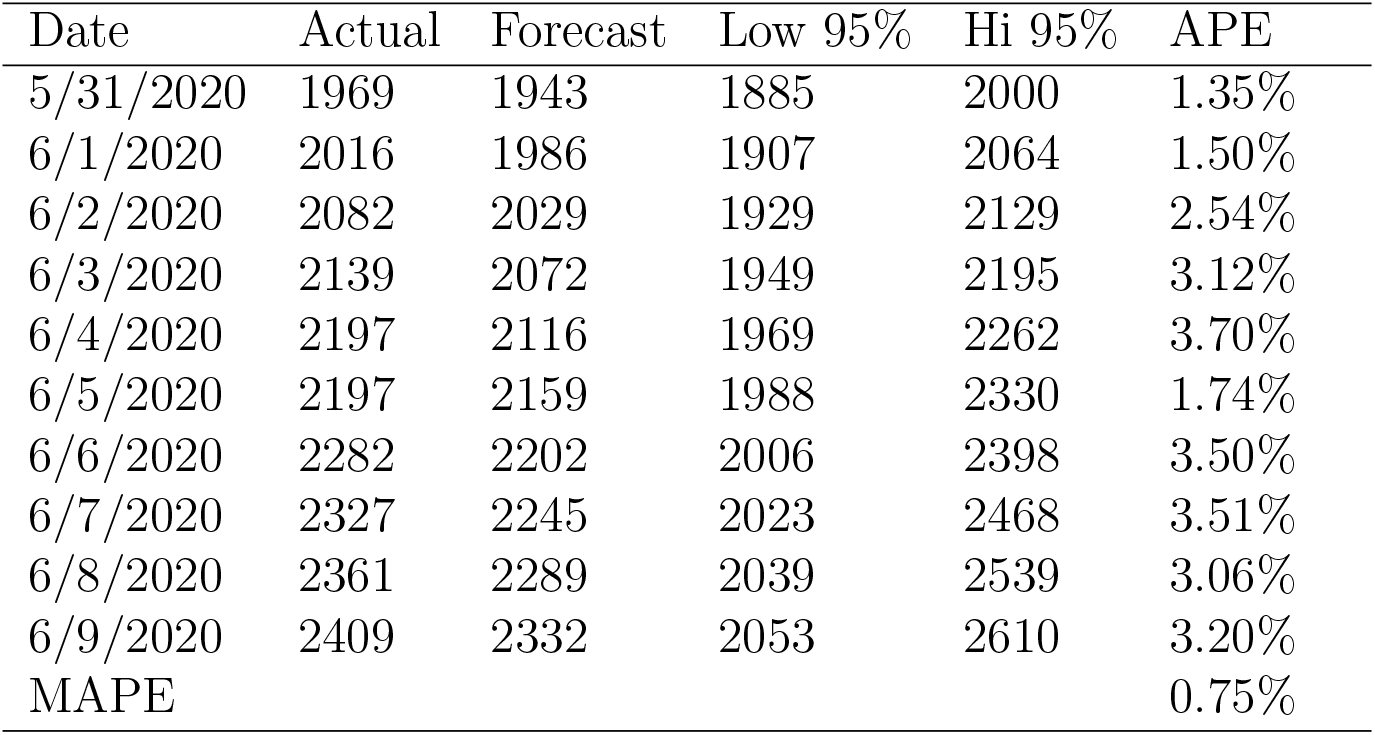
Forecasts, 95% prediction Interval and Absolute percent error.

| Date | Actual | Forecast | Low 95% | Hi 95% | APE |
| --- | --- | --- | --- | --- | --- |
| 5/31/2020 | 1969 | 1943 | 1885 | 2000 | 1.35% |
| 6/1/2020 | 2016 | 1986 | 1907 | 2064 | 1.50% |
| 6/2/2020 | 2082 | 2029 | 1929 | 2129 | 2.54% |
| 6/3/2020 | 2139 | 2072 | 1949 | 2195 | 3.12% |
| 6/4/2020 | 2197 | 2116 | 1969 | 2262 | 3.70% |
| 6/5/2020 | 2197 | 2159 | 1988 | 2330 | 1.74% |
| 6/6/2020 | 2282 | 2202 | 2006 | 2398 | 3.50% |
| 6/7/2020 | 2327 | 2245 | 2023 | 2468 | 3.51% |
| 6/8/2020 | 2361 | 2289 | 2039 | 2539 | 3.06% |
| 6/9/2020 | 2409 | 2332 | 2053 | 2610 | 3.20% |
| MAPE |  |  |  |  | 0.75% |

Fourth strand of forecasts: From June 14 to June 23, 2020. Finally, we updated the data to include the most recent data up until June 10, 2020 and generated 10 days-ahead forecasts. These forecasts with 95% prediction level are portrayed in Fig 2. By June 23, 2020, we forecast that the cumulative confirmed cases will be between 2,730 and 3,263 cases. This implies that there will about 425 new cases for the coming 10 days (June 14 to June 23). There is only 10%, 5% chance that the cumulative confirmed cases will surpass 3,220 and 3,263 cases, respectively by the end of June 23, 2020 while there is also 10%, 5% percent chance that it will be less than 2,677 and 2,634 cases, respectively. Furthermore, We also predict that the total confirmed cases will range from 2,756 to 3,312 by June 28, 2020 with 5 percent of uncertainty.

**Fig 1.**
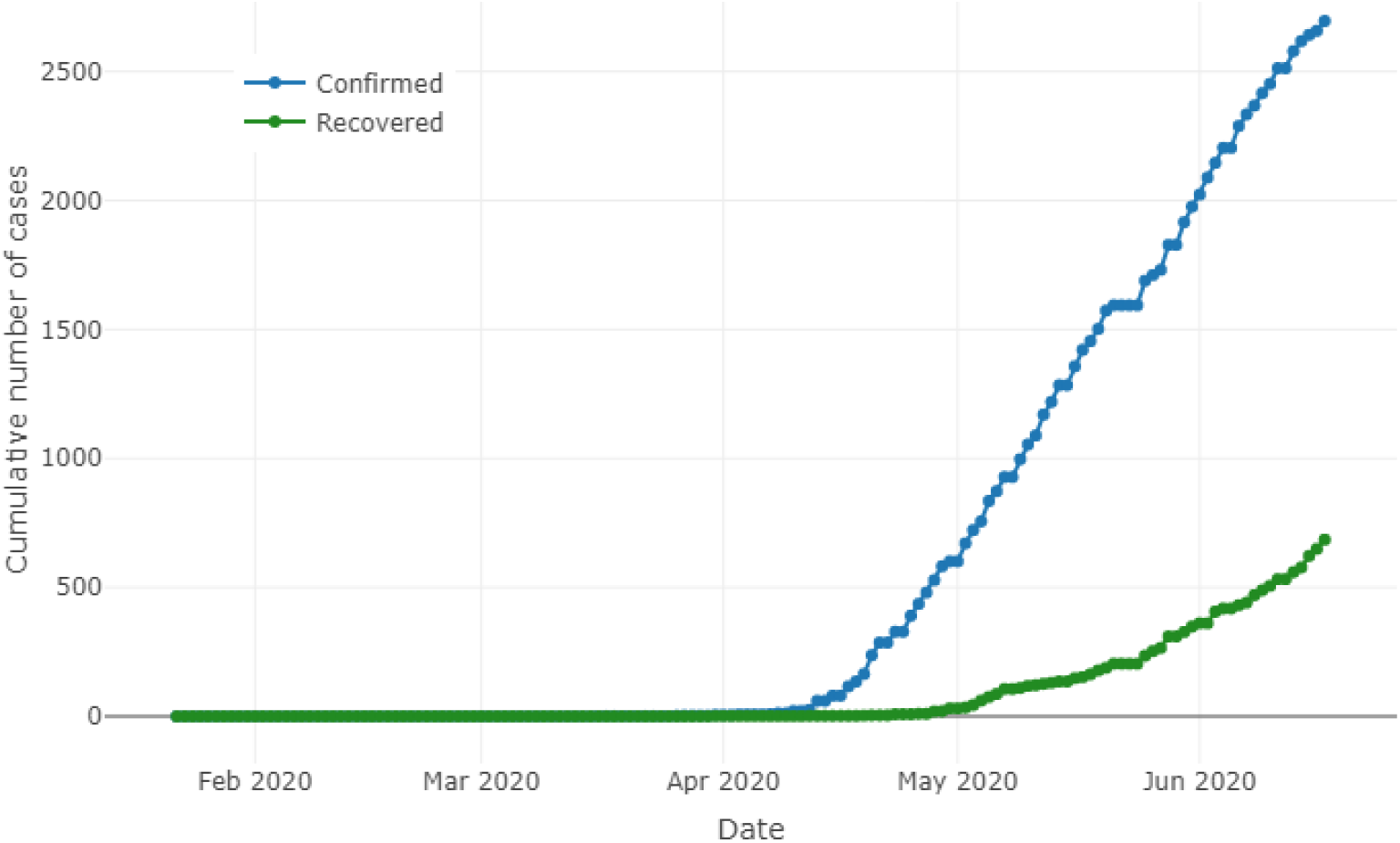
Cumulative confirmed and recovered Covid-19 cases in Somalia.

**Fig 2.**
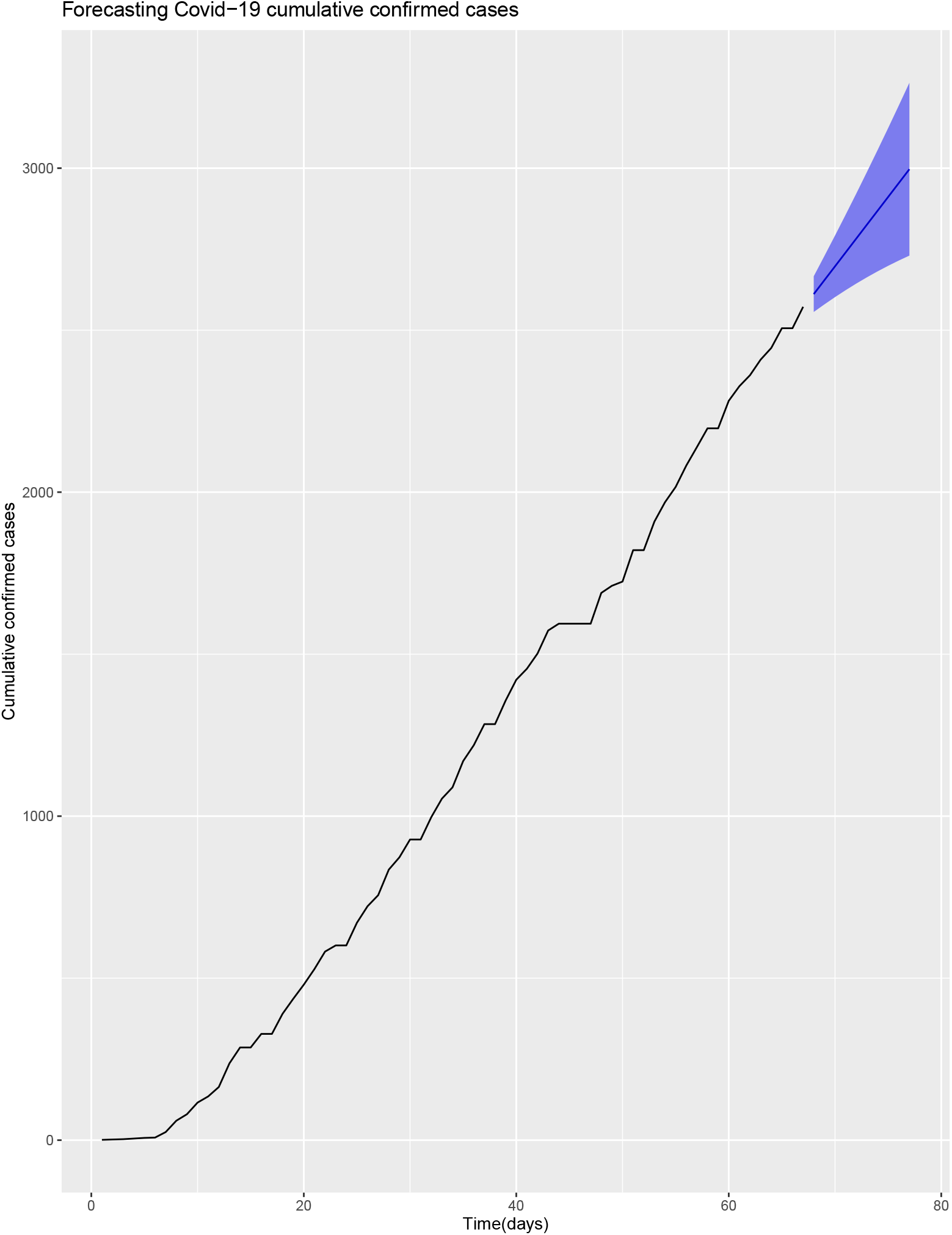
Forecasts with 95% prediction interval.

## Conclusion

To forecast the cumulative confirmed cases in Somalia, we employed an exponential smoothing family especially holt winter’s linear model without seasonality. This model like the rest of forecasting models assume that whatever measures or policies the government took to fight this Covid-19 pandemic in the past will continue to exist in the future. On top of that the accuracy of data used to produce these forecasts is also assumed. In this study, we produced four strands of forecasts and the first three covered the period from May 11 to June 09, 2020. In the first and the third strands of forecasts, the observed and estimated cumulative confirmed cases were near to each other which recorded an average absolute percent error of 0.73 percent and 0.75 percent, respectively. On the contrary, the second set of forecasts recorded an average absolute percent error of 8.7% percent and this implies that the realized and forecasted cumulative confirmed cases were not close as in the first and the third strands of forecasts. The gist of this study was to help the government make necessary plans accordingly. This model provides real time forecasting which can help the government and other stakeholders to get a clear picture on where the maximum number of Covid-19 cases in the country could reach in the future with some uncertainty. With help of these forecasts, the government can make evidence based decisions by utilizing the scarce resource available at its disposal. To this end, forecasting should be the heart and an integral part in the decision making process specifically when the government is dealing with a pandemic like Covid-19. This Covid-19 epidemic did not only pose threat to the public health sector but also severely affected many other sectors such as the economy of the country.

## Data Availability

The dataset used and/or analysed during the current study are available from the Ministry of Health, Somalia.

https://moh.nomadilab.org/

